# Assessing the Quality of Healthcare Practice: A Risk-Based Approach

**DOI:** 10.64898/2026.09.11.26361528

**Authors:** Richard Anthony Despott, Nadine Delicata, Cynthia Mallia, Patricia Vella Bonanno

## Abstract

**Background:** This research article explores a novel framework for assessing the Quality of Healthcare based on principles of Clinical Risk Management, including the development of a theoretical construct for the quality of healthcare services based on stakeholder needs, and a conceptual model for managing risks at an institutional level.

**Methodology:** The study consisted of a cross-sectional secondary assessment of healthcare service and system-level indicators, guided by a structured decision tree based on face, construct, content, and criterion validity. Established performance measures identified from international benchmarking and institutional quality improvement frameworks were classified and validated for assessment of clinical risks according to indicator type.

**Results:** A total of 233 performance indicators were evaluated, of which 200 (86%) consisted of measures for assessing failures in clinical practice. According to the proposed framework, (30%) and (14%) of indicators met the criteria for assessing risks associated with clinical processes and organizational systems design respectively.

**Conclusion:** More targeted use of indicators is required in order to achieve a greater degree of accuracy and actionability of performance assessment measures in healthcare practice. The proposed framework offers an applied method to enhance the specificity of quality indicators and direct efforts to improve systems design and decision-making capability.

## 1. Introduction

The assessment of quality in healthcare is an important aspect of clinical practice as it provides a means to evaluate the capacity to achieve health systems goals, support informed policy decisions and improve the quality of health services. Health regulators and operators in the sector experience significant challenges in evaluating clinical information and translating this into tangible improvements and evidence-based strategies for quality assurance in healthcare delivery [1, 2].

At the level of the service provider, performance assessment involves an evaluation of health care professional (HCP) skills and competencies that are highly complex, which are often confounded by extrinsic aspects of patient care such as teamwork and responsiveness that may not be directly related to health outcomes [3]. Healthcare services are also highly susceptible to the context in which they are provided, as they are impacted by external elements such as organizational systems, financing models and market factors, demographics and developments in biomedical or pharmaceutical technology [4]. Modern concepts of organizational theory such as complexity science and implementation science favour high level strategies underpinned by shared value systems that foster professional networking and interorganizational learning [5]. Such relational forms of governance are the key to developing an over-arching strategy for quality of care that is applicable over a wider scope or generalized institutional setting [6, 7], however this will also require a structure with the oversight and reach to ensure that healthcare organisations employ the necessary resources and methods to support effective quality management systems [8, 9]. Performance assessment and management in healthcare is therefore not only critical to ensure governance and meaningful improvements in service quality but also provides the foundation for regulation of healthcare systems, reducing disparities in standards of practice and avoiding inequities in the quality of care provided to patients [10].

A cohesive framework for benchmarking and sharing of best practice across organizational boundaries demands a clear definition of the standards for managing the quality of healthcare delivery and a set of corresponding measures for assessing performance, which includes indicators for comparing outcomes at a population level and performance measures for evaluating health service quality at an institutional level. Strategies for systems of clinical governance promoted by The Joint Commission on Accreditation of Healthcare Organizations [11] and the World Health Organization include programmes for developing performance measures [12, 13] such as the Health Care Quality Indicators (HCQI) project [14]. Indicators intended for benchmarking of quality at a national or regional health system level generally focus on *outcomes* such as mortality rates to determine what level of performance is achievable compared to other operators in the industry. Performance measures intended to support an internal strategy for quality improvement necessitate a more judicious application of indicators, which reflects the sensitivity and specificity of clinical data, as well as the nature and degree of potential risks that are inherent in the delivery of healthcare services [15, 16]. Indicators intended for improvement of healthcare service quality require performance measures which assess *outputs* in order to understand how internal systems and processes may be enhanced to meet quality objectives more efficiently and more effectively. A risk-based approach may therefore support a more accurate application of quality indicators in healthcare practice, since mechanisms for assessing and improving performance at an institutional level must identify and quantify the underlying failures in order to address them in an adequate and proportionate measure [17, 18].

An effective risk management strategy in healthcare organizations requires a structured framework for clinical governance, whereby performance measures encompass indicators intended to identify potential failures in both the provision of clinical services as well as the operational systems that support these processes [19, 20]. The most widely recognized and established method for auditing health care services and accreditation of healthcare institutions is the model proposed by Avedis Donabedian [21], which focuses on three interconnected components to identify areas for improving quality of care, namely the structures, process, and the outcomes of healthcare delivery **(Figure 1)**.

**Figure 1.**
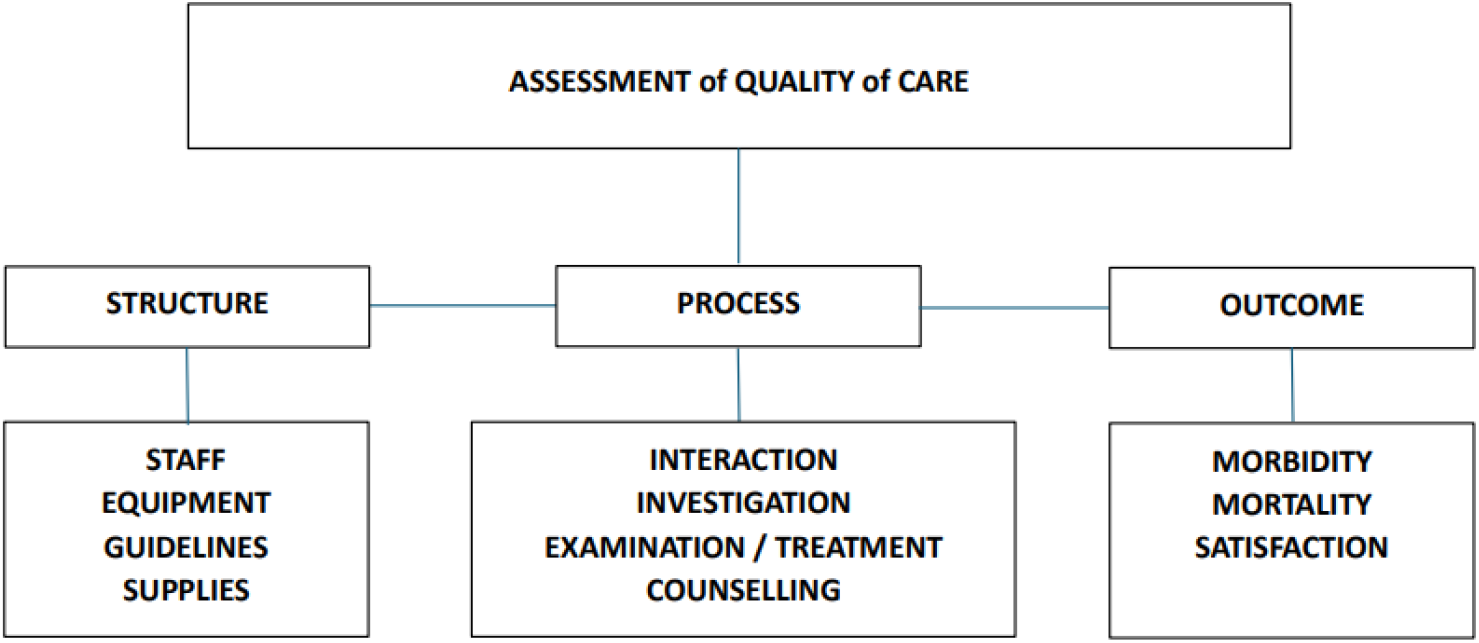
Model for Assessment of Quality of Care ^[21]^. The Donabedian model demonstrates the classification of structure, process, and outcome indicators for assessing the quality of healthcare (1966).

Whilst the Donabedian model provides the foundation for recognising areas of concern in the way healthcare is organized and delivered, a deeper understanding of potential risks in the organizational structures, systems and processes, is necessary to support a more targeted application of performance measures [22] and an effective continuous quality improvement cycle [23, 24]. This is contingent on a standard specification for the quality of care, and an effective suite of performance measures to assess potential risks in an objective, consistent and effective manner [25, 26].

The theoretical basis for the research presented in this article proposes a framework for the assessment of quality in health care grounded in established business management models, which provides the criteria required for validation of performance indicators aimed at managing risk of error in clinical processes and organizational support systems [19, 20]. The validation matrix developed for this research article aligns the types of indicators identified in the Donabedian model with the potential failures encountered at institutional level, linking the assessment and evaluation of clinical data derived from indicators to the development of a more effective strategy for continuous quality improvement and a sustainable approach for achieving higher standards of patient care. This study therefore explores a novel conceptual framework for evaluating the quality of healthcare practices based on the principles of clinical risk management, which includes a theoretical model for identifying the specification for quality of healthcare in terms of stakeholder requirements and a value chain model for validating performance indicators used to assess the conformance of service quality to the desired specification.

## 2. A Framework for Evaluating Quality in Healthcare

A framework for evaluating the quality of healthcare was developed using tools established in business process administration to ensure consistency in purpose and function between the construct of health care quality and the performance measures intended for assessing and managing clinical services. This included a stakeholder matrix to establish the design quality of the service (the degree with which the specification meets customer needs), and a value-chain model to identify potential failures in the quality of conformance (the degree with which a service meets the specification) [27].

### 2.1 Theoretical Construct for defining the Quality of Healthcare

The conceptual definition for quality of healthcare is described by the degree to which health services for individuals and populations increase the likelihood of desired health outcomes [28, 29]. The specification of design quality must therefore reflect the needs of key stakeholder groups [30, 31], in order to ensure that indicators employed as a formative mechanism for service improvement assess the relevant dimensions of quality [32, 33]. Health outcomes driven by stakeholder value may broadly be divided into the core attributes that are directly associated with clinical services and aspects of quality related to health systems performance [34]. These may be differentiated further into dimensions of intrinsic quality which directly impact health benefits, and dimensions of extrinsic quality related to client experiences and expectations [35, 36]. The strategic domains for quality of care within this construct are classified according to the key stakeholder groups in **Figure 2**.

**Figure 2.**
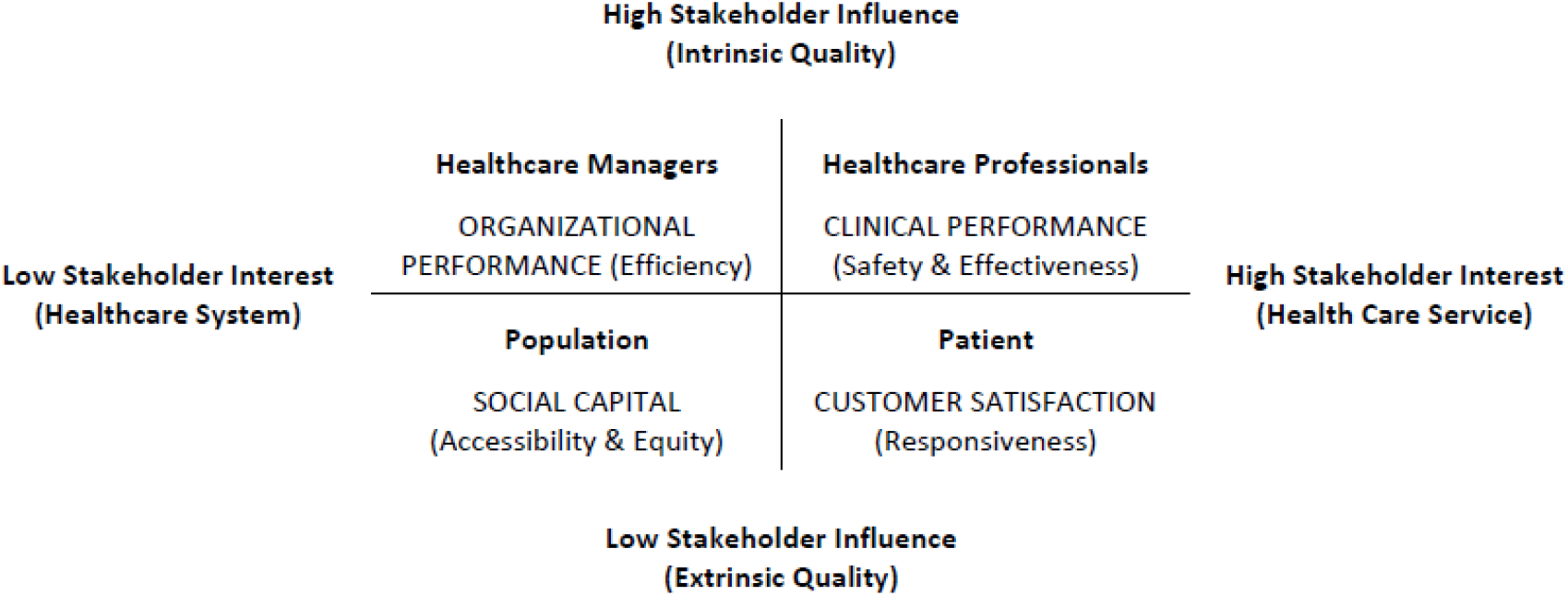
Stakeholder Model for Quality of Healthcare Practice ^[37]^. The Stakeholder model demonstrates the relationship between the strategic domains of quality in healthcare, based on the classification by author (2025).

The stakeholder model described above indicates that healthcare systems quality at a population level is primarily concerned with benchmarking indicators for comparison of equitable outcomes such as morbidity and mortality. At an institutional level, performance measures intended for quality improvement purposes require structural indicators that bear insight into the quality of organizational performance e.g. staff to patient ratios, and process indicators for managing service quality at a transactional level including customer satisfaction e.g. waiting times and compliance with clinical standards [38, 39].

### 2.2 Conceptual Model for assessing the Quality of Healthcare

According to the strategic domains identified in the Stakeholder model above, an effective evaluation of the dimensions of quality cannot be assured if performance indicators are applied arbitrarily or summarily grouped to address a composite value [40]. Assessment of potential failures at an institutional level must contend with different types of clinical risks which involve the effectiveness and safety of clinical services, as well as potential failures in the performance of organizational support systems. The scope of performance measures with regard to clinical risk management may be described by the cumulative act effect theory (Swiss Cheese model), which defines potential errors as a product of first-order risks or active errors that are inherently associated with the behaviour of frontline workers in direct contact with patients, and second-order risks or latent errors in the systematic processes that allow active errors to cause harm [41]. The distinction between risks associated with clinical services provided directly by health care professionals and systemic risks that arise from support activities managed by the administrative structure is illustrated in **Figure 3**.

**Figure 3.**
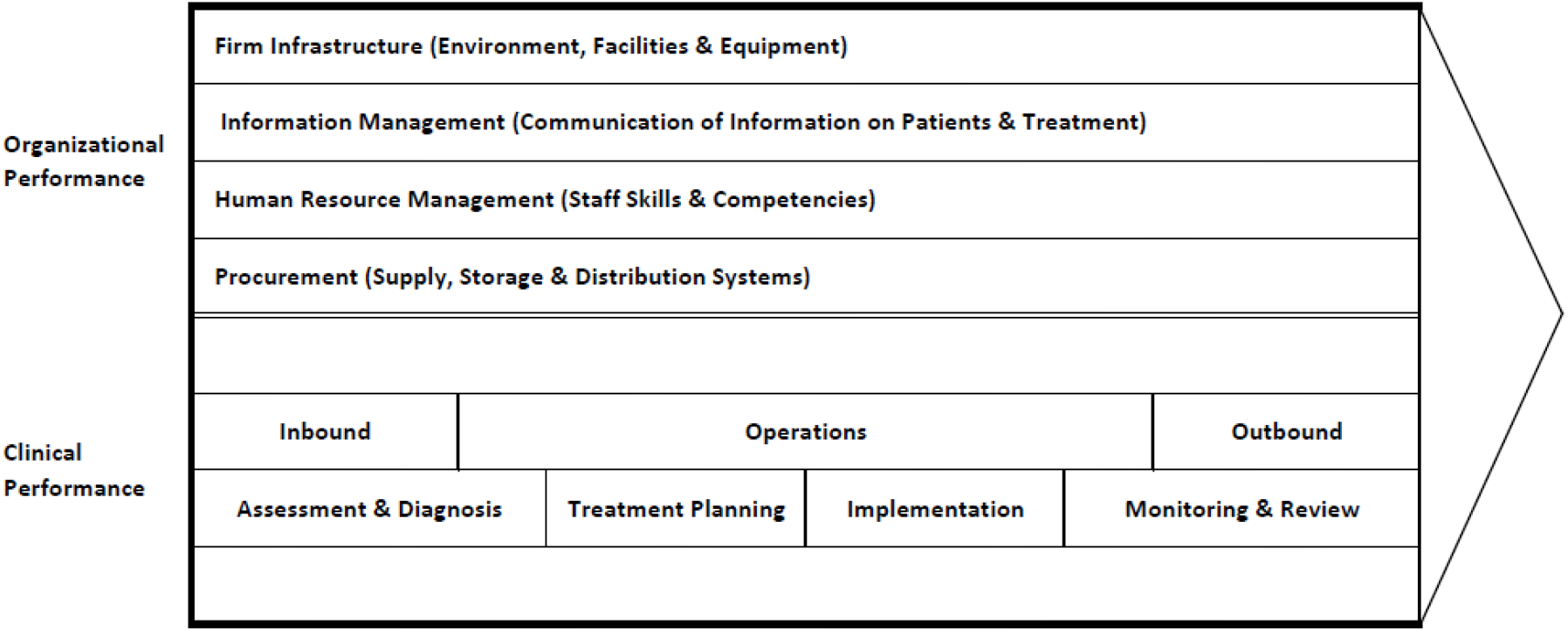
Value Chain Model for Clinical Risk Management (adapted from Porter ^[42]^) The Value Chain Model demonstrates the relationship between the primary activities and the support activities in healthcare organizations.

The Value Chain model shown above provides a generic framework for assessing intrinsic quality based on the commonality of organizational structures, systems and processes within a hospital or healthcare institution [42, 43]. Clinical activities involve a series of interconnected and iterative, yet distinct decision-making processes by health care professionals, each of which represents a critical control point for managing preventable harm to patients due to medical errors [44, 45]. In this case, the intrinsic quality afforded by a clinical decision is defined in terms of failures in conformance to best practice since ‘added value’ is a return to normality or optimal health [46, 47]. The quality of clinical performance, corresponding to the dimensions of intrinsic quality (safety and effectiveness), is therefore a function of the risk of medical error associated with a treatment process, which may be calculated in relative terms as a product of the frequency and severity of patient harm [37, 48]. This means that indicators intended for managing the quality of clinical care should also be prioritized by considering the relative risk in terms of the occurrence and the potential for harm [49, 50]. A more accurate assessment of clinical risk is therefore proposed by applying structural indicators to identify latent errors in the design quality of organizational structures and systems, and process measures to assess the risk magnitude of medical errors in healthcare services.

## 3. Method

The research consisted of the classification and validation of performance measures to identify inconsistencies in the types of indicators used for assessing quality at institutional level, based on the potential risks that are inherent in the delivery of healthcare. This was achieved through a structured decision tree matrix designed to evaluate the face, construct, content and criterion validity criteria established by the theoretical models described in the previous sections **(Table 1)**.

**Table 1.** Validation matrix for Indicators of Quality in Healthcare delivery (developed by author) Table 1 shows the criteria used to identify and validate performance indicators intended to measure the quality of care in healthcare practice, with relevance to the type of potential risks that are inherent in clinical processes and support activities carried out at an organizational level.

|  |  |  |
| --- | --- | --- |
| **Face Validity** |  |  |
| Quality Indicator | Health Care Service | The indicator measures the quality of a health care professional service |
|  | Healthcare System | The indicator measures a benchmark of the healthcare delivery system |
| **Construct Validity** |  |  |
| Service Quality | Intrinsic | The indicator measures the quality of a medical service |
|  | Extrinsic | The indicator does not measure the quality of a medical service |
| **Content Validity** |  |  |
| Strategic Domain | Clinical Performance | The indicator is a measure of Safety & Effectiveness provided by a health care service |
|  | Customer Satisfaction | The indicator is a measure of Responsiveness provided by a health care service |
|  | Organizational Performance | The indicator is a measure of Efficiency provided by a healthcare system |
|  | Social Capital | The indicator is a measure of Equity & Accessibility provided by a healthcare system |
| **Criterion Validity** |  |  |
| Type of Indicator | Outcome Indicator | The indicator is a measure of Morbidity, Mortality and Customer Satisfaction |
|  | Structure Indicator | The indicator is a measure of Staffing, Equipment, Guidelines and Supplies |
|  | Process Indicator | The indicator is a measure of Interaction, Investigation, Treatment and Counselling |

The criteria listed in Table 1 differentiate indicators of health service quality from benchmarking measures and indicators of extrinsic quality, which fall outside the scope of clinical risk management. The validation of performance indicators was assessed according to the types of measures established in the Donabedian model and the criterion validity for evaluation of clinical risks, whereby first order risks due to human error (medical error) are identified through process indicators whilst second order risks due to latent error in system design are identified through structural indicators. The classification and validation of individual indicators according to face, construct, content and criterion validity was performed and moderated independently by the authors of this article using the validation matrix as a focal point for consensus in case of ambiguity or duplicity of the measures.

The scope of the research considered indicators and performance measures pooled from indicator sets designed for benchmarking and quality improvement, which were identified from the tools available in the literature **(Table 2)**. Indicator sets that were not exclusively concerned with the dimensions of healthcare, such as guidelines developed by the National Institute for Health and Care Excellence [51], tools intended for a defined patient population or health system [52, 53], indicators limited to a specific dimension of quality [54], and tools designed for a particular scope such as application to the European Reference Network or health funding program [55, 56] were omitted from the scope of the study. The tools selected for this study included the HealthCare Quality and Outcomes (HCQO) indicators established by the Organisation for Economic Cooperation and Development [13], and the Performance Assessment Tool for Quality Improvement in Hospitals (PATH) established by the World Health Organization [57]. These sets were preferred due to their design as the most representative tools for comparison of national data and quality improvement respectively. Ethical approval was not required because the data did not involve any specific reference to human subjects or identifiable patient data, and the individual indicators considered for the study were publicly available in the literature.

**Table 2.** Examples of Indicator sets for Assessment of Quality in Healthcare. Table 2 shows examples of indicator sets that are intended for international benchmarking and institutional quality improvement frameworks that are publicly available in the literature.

| Indicator Set | Description |
| --- | --- |
| National Institute for Health and Care Excellence Indicator Set <sup>[51]</sup> | Menu of indicators designed to help define and measure quality in health, public health and social care sector. |
| Health System Performance Assessment Tool (2015) <sup>[52]</sup> | A set of 57 indicators selected for a report on the Performance of the Maltese Health System |
| Quality Indicators of inpatient paediatric and newborn care (2023) <sup>[53]</sup> | A tool for prioritising WHO indicators for paediatric and neonatal quality indicators in hospitals. |
| Europe European Society for Quality in Healthcare safety indicators (2007) <sup>[54]</sup> | Assessment Framework Approach to develop an internal indicator set for EU patient safety indicators (SImpatIE) |
| European Reference Network Indicators (2019) <sup>[55]</sup> | Self-Assessment tool of operational criteria created by the European Commission for Healthcare Providers to integrate healthcare systems |
| World Health Organization Quality of care indicators (2024) <sup>[56]</sup> | Quality of care indicators to support implementation of the Health Quality Fund in Romania |
| Organization for Economic Cooperation and Development (2024-2025) <sup>[13]</sup> | HealthCare Quality and Outcomes (HCQO) list of indicators for data collection |
| World Health Organization (2007) <sup>[57]</sup> | Performance assessment tool for quality in hospitals (PATH), issued by the WHO Regional Office for Europe |
| European Core Health Indicators <sup>[58]</sup> | Data Tool for comparable health information developed by the European Commission. |
| Australian Commission on Safety and Quality in Health Care. <sup>[59]</sup> | Hospital indicators identified by the commission for clinical risk mitigation strategies |
| WHO Global Reference list of Core Health Indicators (2015) <sup>[60]</sup> | A general reference and guide for standard indicators and definitions for monitoring health situations |

## 4. Results

The study considered a total of 233 performance measures that were included in the HealthCare Quality and Outcomes (HCQO) indicators and the Performance Assessment Tool for Quality Improvement in Hospitals (PATH) indicator sets. The complete classification of indicators by the validation criteria according to the strategic domains in the Stakeholder Model are presented as a supplementary appendix to the manuscript, which is summarized in **Figure 4** below.

**Figure 4.**
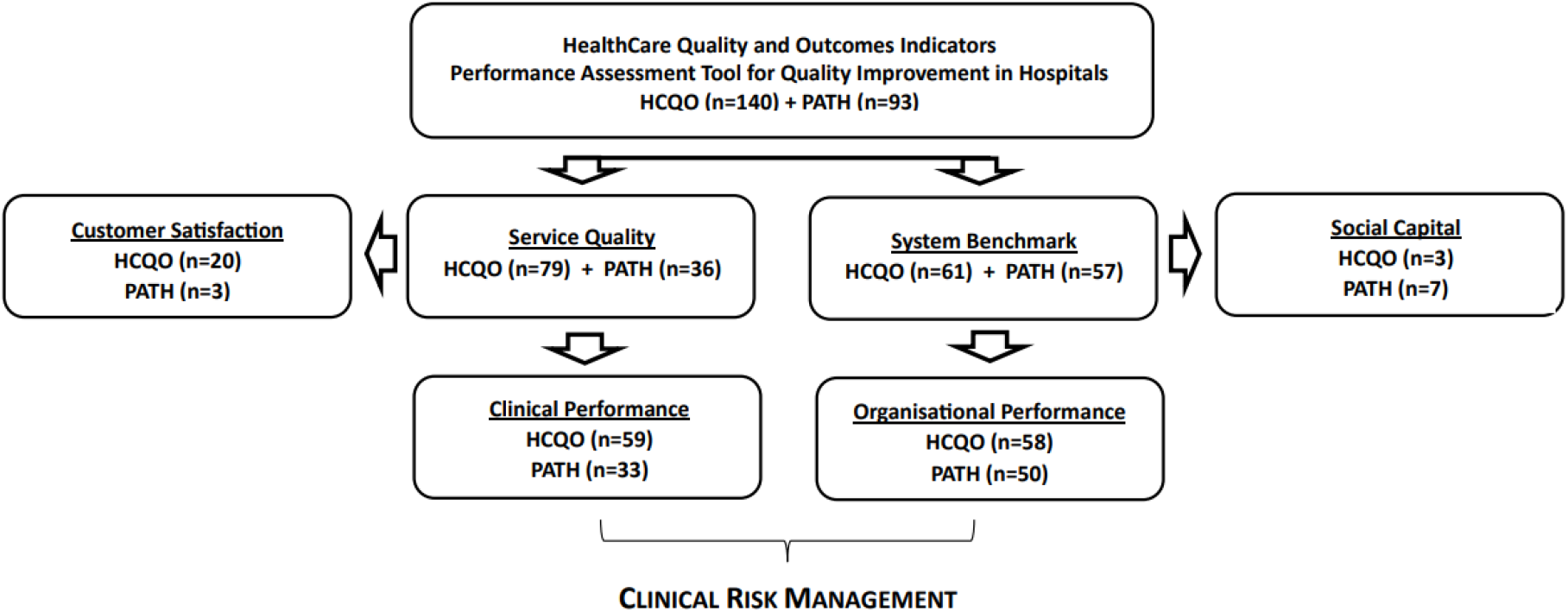
Classification of Healthcare Quality Indicators with relevance to Strategic Domain (n=233) Figure 4 shows the classification of indicators according to the strategic domains identified in the Stakeholder Model for quality of healthcare practice

According to the decision tree matrix described in Table 1, approximately 49% (79 HCQO + 36 PATH) of indicators were classified as measures for assessing healthcare services. The majority (80%) of healthcare services indicators related to the quality (safety and effectiveness) of clinical performance (59 HCQO + 33 PATH), whilst the majority (92%) of health systems indicators focussed on the quality (efficiency) of organizational activities (58 HCQO + 50 PATH). The number of indicators for each of the strategic quality domains with relevance to the type of measure (structure, process, outcome) required to assess specific clinical risks described in the Value Chain Model is illustrated in **Figure 5**.

**Figure 5.**
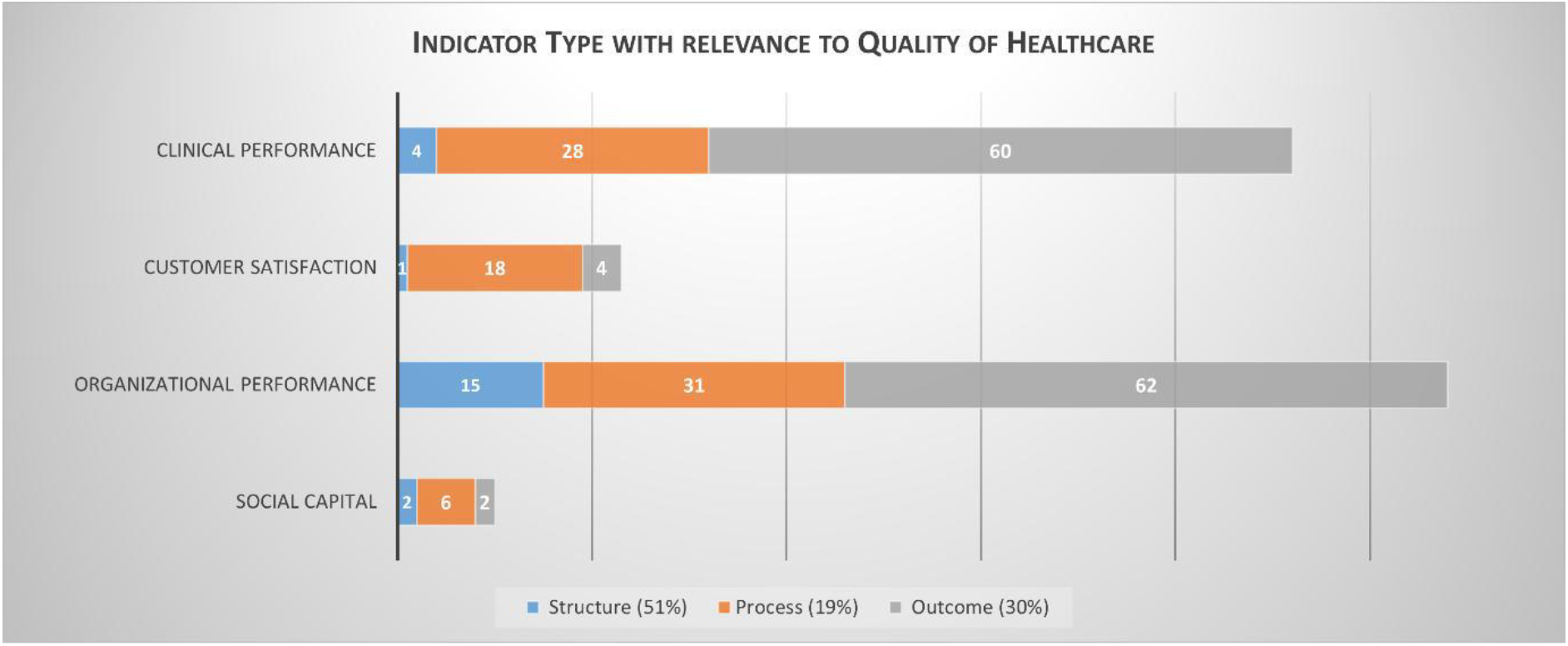
Validation of performance indicators with relevance to Quality of Healthcare (n=233) Figure 5 shows the type of indicators according to the strategic domains identified in the Stakeholder Model for quality of healthcare practice

## 5. Theoretical Framework for Validation of Healthcare Quality Indicators

The framework models described in the methods section, namely the Stakeholder Matrix (Figure 2) and the Value Chain model for assessment of quality in healthcare (Figure 3), provided the theoretical basis for the validation of indicators according to their strategic value. Whilst outcome indicators offer a rational measure for benchmarking extrinsic aspects of quality, a clinical risk management approach re-directs efforts for quality improvement in healthcare towards the safety, effectiveness and efficiency of patient treatment and support activities carried out at an institutional level [61, 62].

### 5.1 Implications for Assessment of Quality in Healthcare

The Stakeholder Model for Quality in Healthcare Practice (**Figure 2**) provides a conceptual definition that reconciles different dimensions in the design quality of healthcare under a single construct and specifies the criteria against which performance must be evaluated in order to identify areas for improvement within a particular strategic domain [37]. This is consistent with previous studies, which distinguish between the quality of health systems performance and health care services [19, 20] and differentiate between assessment of intrinsic quality identified with patient safety and effectiveness and extrinsic quality based on client perceptions [36, 63]. The value-chain model (**Figure 3**) explores the strategic domains relevant to clinical risk management and identifies the criterion validity of performance indicators which are intended for assessing quality and promoting systematic improvement at the level of health services providers [64, 65]. Structural indicators, therefore, provide more effective measures for targeting potential risks of latent error associated with deficiencies in the design of organizational structures and support systems, whereas process-based indicators provide a more accurate evaluation of conformance to good practice, which may give rise to medical errors due to inherent risk of human failure in clinical-decision making processes [66, 67].

The Value Chain Model for Assessment of Quality in Healthcare Services further demonstrates how performance indicators have a distinct role in promoting a cohesive strategy for clinical risk management, by facilitating a data driven approach to improvements in service quality [68, 69]. This is achieved by providing a strategic basis for ensuring that the ‘hard’ organizational elements, which can more easily be defined and controlled, foster and support the management of the less tangible ‘soft elements’ that impact organizational culture and shared values [70]. Medical errors of diagnosis and treatment planning are essentially design errors which should be managed through decision support systems, including for example drug information services and medication review by clinical pharmacists. Medical errors associated with the implementation and execution of the treatment plan ultimately provide a key yardstick for managing safety and effectiveness of patient care through more effective training programs for frontline carers and improving in-process quality control in commensurate measure with the risk magnitude of the clinical decision-process [37, 48].

### 5.2 Implications for Clinical Risk Management

The design quality for the indicator sets compiled by expert groups showed a high degree of selectivity for the assessment of intrinsic quality (84% and 89% for HCQO and PATH indicators respectively) and maintained a fair balance between measures of clinical and organizational performance (**Figure 4**). The indicators for domains of intrinsic quality of care however exhibited a lower degree of specificity in terms of criterion validity for clinical risk management, namely process indicators (30%) for measures of clinical performance and structural indicators (14%) for measures of organizational performance, which therefore may not represent a complete or accurate assessment of the underlying failures for the scope of quality improvement at institutional level (**Figure 5**).

The validation criteria developed in this research article identified a number of gaps and potential inconsistencies when applying indicators for assessing healthcare performance, which imply that providers are limited in translating the information from indicator sets into meaningful operational strategies. The majority of indicators aimed at assessing clinical effectiveness and safety consisted of outcome measures (65%), including for example re-admission rates after surgery and risk-based mortality (PATH), pressure ulcer prevalence and healthcare associated infections (HCQO). Whilst outcome measures by definition lack the detail on the underlying causes that must be addressed to improve service quality, these metrics are also limited with regard to benchmarking since the comparison of performance must take into context the impact of organizational differences and the institutional setting. Thus, whilst outcome indicators provide a general benchmark for the areas of practice where clinical outcomes can be improved, they do not distinguish between failures in clinical practices and organizational systems, for example higher re-admission rates or healthcare associated infections are not necessarily indicators of clinical safety and effectiveness if the setting is a geriatric or oncology facility, or if there is a staff shortage or inadequate maintenance of ventilation and air quality in the operating theatre. Similarly, structural indicators such as volume of prescribed antibiotics are not specific enough to assess the quality of clinical performance, since the evaluation of safety and effectiveness requires measures to identify failures in the decision-making processes (medical error). Measures of organizational performance largely consisted of process indicators such as waiting times and length of hospital stay (29%) and outcome indicators of mortality and morbidity (57%), however if these are to be applied to assess operational efficiency structural aspects such as staffing ratios and availability of medication and medical equipment must also be considered. The overriding application of outcome indicators in areas such as acute care, integrated care, end of life and cancer care cannot ultimately be interpreted as a definitive measure of failure in organizational performance if the structural components are adequate for the demands of the patient groups.

It is noteworthy that whilst the overall number of indicators for clinical risk management pooled from the selected indicator sets was relatively high (89%), indicators for assessing intrinsic quality of clinical services (safety and effectiveness) was significantly lower (40%), and the number of these measures meeting the criterion validity to detect potential risk of errors was less (12%), which indicated a low propensity for improving the quality of patient care. The highest degree of validation was recorded for performance measures of customer satisfaction (78%) which reflects the relative difficulty encountered when transposing traditional methods for performance assessment to clinical services and substantiates the argument that a framework for aligning indicators with the evaluation of potential risks can support a more accurate and better management of healthcare practice.

## 6. Limitations and Future Research

The theoretical models described in this article qualify the application of healthcare quality indicators in terms of managing stakeholder demand (**Figure 2**) and the potential risks encountered in the delivery of healthcare (**Figure 3**). The logical design presents a structured framework for more accurate assessment of failures in design quality and conformance to standards of clinical practice however, risk analysis introduces inherent limitations due to the subjectivity of the assessor and the degree of overlap of contributing factors to health indicators. The degree of impact on the results presented in this paper was moderated through the creation of a validation matrix, however this limitation should not in any case detract from the value of the theoretical framework linking the audit of clinical information with risk control measures more effectively and reducing the occurrence and cost of adverse events more efficiently based on the relative risk of preventable harm [67, 71]. Another limitation of this study is the selection of just two indicator sets which may not provide a broad representative sample of the instruments developed for such purposes or sufficient detailed insight into the existing gaps for assessment of quality, however the preliminary results provide a clear indication that accuracy can be improved significantly by the proposed method.

The alignment of established indicator types with the evaluation of strategic domains provides an opportunity to target data collection more effectively depending on whether clinical risk management strategies are aimed at improved systems design or quality control of treatment processes [72, 73]. Advancing the understanding of factors impacting clinician decision-making in complex hospital environments also avoids the subjectivity and imprecision of quality improvement efforts based on social constructs and creates a positive feedback loop which dispels blame culture and sustains a long-term strategy for organizational learning and patient safety culture [74, 75]. Developing performance indicators which target the systematic identification and evaluation of clinical risk factors is therefore a critical success factor for leveraging the untapped potential of real-world information and promoting continuous improvement.

The application of the theoretical framework to published indicator sets requires further research to determine whether a risk-based approach can achieve meaningful results in a real-world setting. This includes studies which validate the potential for clinical risk management to reduce the incidence of medication errors in clinical practice [76], as well as a better appreciation of how data-driven methods can help overcome long-standing difficulties for transferring quality management methods to the healthcare environment. At population level, more insight is needed on the feasibility of applying risk-based performance measures to support better regulation and sharing of best practice across different healthcare settings [77].

## 7. Conclusion

This article explores an original framework for assessing quality in healthcare based on principles of risk management, which adapts the types of indicators described in Donabedian’s model with the analysis of failures in clinical practice to realise a more accurate understanding of the solutions required to improve service quality. The theoretical basis for the study included the identification of specific dimensions associated with the strategic outcomes for quality of care based on stakeholder requirements and the development of a value-based model to target the root causes underlying failures in the delivery of healthcare. The research differentiates between *outcome indicators* used for benchmarking of healthcare systems at a population or national level and performance indicators intended for assessment of service quality. A more accurate evaluation of performance for quality improvement purposes is proposed by applying *process indicators* to identify failures of conformance in clinical practice (active errors), and *structural indicators* to identify failures in the design quality of institutional structure and systems (latent errors). The framework presented in this article does not presuppose that the presence of quality cannot be measured or improved to all extents possible by the indicator sets already identified in the literature by expert panels however it offers a practical method to increase the accuracy of tools applied in assessing quality and improve actionability by concentrating on identifying and quantifying the underlying risk of failure.

The risk management approach focuses on the identification of opportunities to reduce patient harm and improve service outcomes within a specific healthcare process, whilst also supporting quality improvement systems and organizational efficiency through the development of cohesive, data-driven strategies which target both the inherent risk of preventable harm due to human error and the systematic factors that influence interventions made by health care professionals. In addition, the use of quality assurance methods using root cause analysis are a critical success factor to discourage blame seeking culture because they link the assessment of clinical practices with equitable standards in the quality of care and direct efforts towards areas of practice with high risk of preventable harm. It is noteworthy that this method does not restrict the definition of an error with a particular health care profession but focuses on the type of failure associated with the clinical decision-making process so that performance assessment retains a focus on root cause analysis and continuous improvement. This proposed framework for clinical risk assessment is ultimately based on generic structures, systems and processes, which means it can therefore be applied on a broad scale independently of any clinical specialty, healthcare setting or organizational structure. The implications for the evolution of assessment methodologies based on analyses of data impacting clinical decision-making cannot be understated, since the accuracy and effectiveness of clinical audit is the key to promoting a sustainable patient safety culture and achieving higher standards of healthcare service quality.

## Data Availability

All data produced in the present study are available upon reasonable request to the authors

